# Socioeconomic risk factors and human immunosuppressive conditions are major drivers of human leishmaniasis in southern Europe

**DOI:** 10.64898/2026.03.16.26348465

**Authors:** Danyang Wang, Kevin D. Matson, Anouschka R. Hof, Eduardo Berriatua, Carla Maia, Federica Bruno, Germano Castelli, Pedro Pérez Cutillas, Jose Risueño Iranzo, Elena Verdú Serrano, Fabrizio Vitale, Ângela Cristina Gomes Xufre, Frank van Langevelde

## Abstract

**Background:** Leishmaniasis is endemic in southern Europe with high risks of outbreaks and geographical spread. However, the risk factors for human leishmaniasis are understudied in this region.

**Methodology:** To evaluate these risk factors, we tested associations between leishmaniasis incidence and an array of variables related to socioeconomics, immunocompetence, climate, land use, biodiversity, and ecology (i.e., the pathogen and its vectors and reservoirs).

**Findings:** Socioeconomic factors, such as demography, occupation, and housing conditions, were strongly associated with leishmaniasis incidence in endemic regions. The specific factors and the magnitude of their impacts varied among the five countries studied. Human immunosuppressive condition was highly correlated with leishmaniasis risk in Spain and Italy. Climate likely delineated leishmaniasis-free regions from endemic regions in France. Our results suggest that climate change alone may not drive the spread of leishmaniasis within this century. Pathogen hazard and reservoir abundance affected leishmaniasis risk more than vector hazard in countries where data were available. Biodiversity was weakly, negatively related to leishmaniasis.

**Significance:** Our results highlight the importance of socioeconomic risk factors and immunosuppression for human leishmaniasis, suggesting potential implications for disease control and prevention policies. Surveillance of *Leishmania* spp. in humans, vectors, and reservoirs; assessment of reservoir abundance; and data accessibility are crucial for disease prevention and preparedness. Because of possible biodiversity regulation, efforts to understand and control leishmaniasis could benefit from a One Health approach that involves epidemiologists, social scientists, and ecologists, among others.

**Author summary:** Leishmaniasis is a common disease in tropical and subtropical regions, but it also occurs in southern Europe. While some infectious disease experts are concerned that climate change might lead to the emergence of new diseases in new areas, the main factors shaping leishmaniasis in Europe are not well understood. This study considered a wide array of potential risk factors in a variety of categories, ranging from climate and nature to how people live and work. We found that the risk for leishmaniasis is mostly related to occupation, housing type, age, and sex, though the exact risk factors change from country to country. People with weakened immune systems face high risk, and infected animals pose a major threats, but climate change alone may not drive the spread of leishmaniasis as much as once feared. Interestingly, a healthy variety of wildlife may help keeping the disease in check. To limit the impacts of leishmaniasis, we need to protect the most vulnerable populations, such as people living with weakened immune systems, working in high-risk sectors, residing in single-dwelling buildings, or experiencing or facing homelessness. We need to monitor the parasite in people and animals and share those surveillance data openly. Ultimately, we need a “One Health” approach where doctors, social workers, and scientists work together to keep our ecosystems healthy and our communities safe.

## 1. Introduction

Leishmaniasis is a neglected tropical disease with high burdens. (1) In Europe, leishmaniasis is endemic in the Mediterranean region with high risks of outbreaks and geographical spread. (2) The causative agent of leishmaniasis is the protozoan parasite *Leishmania.* Several *Leishmania* species occur in and around Europe: *Leishmania infantum* in southern Europe, Turkey, North Africa, and the Middle East; *L. major* in North Africa, the Middle East, and Turkey; *L. tropica* in North Africa, the Middle East, Turkey and Greece (sporadic); *L. donovani* in Turkey, the Middle East and Cyprus (sporadic).(2) The lifecycle of *Leishmania* species requires a vertebrate host and an insect vector (i.e., Phlebotomine sand fly (Diptera: Psychodidae)) for transmission. (2) *L. infantum* and *L. major* can be transmitted from animal to human (i.e., zoonotic), whereas *L. donovani* is transmitted between humans (i.e., anthroponotic). (2) *L. tropica* can be transmitted through both pathways. (2) In the Mediterranean basin where *L. infantum* is widespread, domesticated dogs (*Canis familiaris*) are the main domestic reservoirs, whereas wildlife, such as Iberian hares (*Lepus granatensis*), can act as sylvatic reservoirs. (2) Risk factor studies of leishmaniasis were thus far mostly at small spatial scales (i.e., up to national level). (3) These studies revealed that the risk factors of leishmaniasis are multidimensional. (3) However, due to the small scales and the often location-specific variables in these studies, it is hard to compare the relative importance and the transferability of the identified risk factors. Recently, a large-scale study in Europe focused on climatic variables explaining leishmaniasis and found that climate change possibly facilitated the northwards spread of leishmaniasis. (4) However, understanding the full spectrum of disease risk factors at large scale is essential for optimizing disease control and prevention policies. (5)

Disease risk can be measured via hazard, exposure, and vulnerability. (6) Risk is defined as the likelihood of adverse events, hazard is a potential source of harm, exposure is the likelihood of contact with the hazard, and vulnerability is the possibility, given exposure, that the hazard can cause harm. (6)

Risk = Hazard × (Vulnerability × Exposure) (6)

This formula can be applied to leishmaniasis: the number of new cases in a population per unit of time = the potential presence of infected vectors × (the probability of morbidity upon exposure × the likelihood of contact between humans and infected vectors).

Within the geographic areas where the pathogen (i.e., pathogen hazard) and vectors (i.e., vector hazard) co-occur, climatic, land-use, and biodiversity variables can modulate pathogen-vector interactions and thereby influence transmission cycles. (7) Contact rates are influenced by land-use and socioeconomic factors, such as housing conditions, occupation, human demography (e.g., sex ratio), education level, and human population density. (3) Given contact of humans with infected vectors, morbidity is related with human immunocompetence (e.g., immunodeficiency due to human immunodeficiency virus (HIV), cancer and immunosuppressive treatments) and socioeconomic factors, including human demography (i.e., age), socioeconomic status, and health services. (3)

In this study, we tested the associations and their strengths between human leishmaniasis incidence and vector habitat suitability, *Leishmania* infection in vectors and animal reservoirs, climate, land use, biodiversity, human immunosuppressive conditions, and socioeconomic variables. This allowed us to rank the risk factors of leishmaniasis in southern Europe. Whenever possible, we made predictions about leishmaniasis in the future.

## 2. Method

### 2.1 Leishmaniasis incidence and standardized incidence rate

Leishmaniasis incidence data were hospital admissions collected from the Ministry of Health in Spain (https://www.sanidad.gob.es/en/estadEstudios/estadisticas/cmbdhome.htm), supplemented by reported cases from Portugal, France, Italy, and Greece (Table 1). (8) All data were anonymized and spatially aggregated to nomenclature of territorial units for statistics (NUTS) 2024 level 2 (regional policies) or 3 (local specifications). (9)

**Table 1.** Overview of leishmaniasis data.

| Country | Response variable | Source | Spatial resolution | Period | Sex/ Age | Risk | Sample size (NUTS) | Regression model |
| --- | --- | --- | --- | --- | --- | --- | --- | --- |
| Portugal | Incidence | Literature | NUTS 3 | 2010-2020 | No | Absolute | 23 | Poisson |
| Spain | SIR | Ministry of Health | NUTS 3 | 1997-2015 | Yes | Relative | 52 | Poisson |
| France | Incidence | Literature | NUTS 3 | 1999-2012 | No | Absolute | 96 | Zero-altered Poisson |
| Italy | Incidence | Literature | NUTS 2 | 2011-2016 | No | Absolute | 21 | Poisson |
| Greece | Incidence | Literature | NUTS 3 | 2009-2018 | No | Absolute | 52 | Poisson |

SIR: standardized incidence ratio; NUTS: nomenclature of territorial units for statistics

Spanish datasets contained information on sex and age; published data from the other countries did not have these attributes. Thus, for Spain we could compute the standardized incidence ratio (SIR) to model the relative risk, allowing for comparison among populations with different sex-age structures by controlling for demographic variation. (10) SIR is the quotient of observed and expected numbers of cases: a value > 1 indicates higher risk areas where more cases were observed than expected; a value < 1, the opposite. (11) The expected cases reflect the situation where a specific population has the same sex and age structure as the national population. (11) All computations were conducted in RStudio (version 2024.12.0+467). (12) Major functions and packages used are listed in supporting information (S) 1.

### 2.2. Predictor variables

We tested 400-700 predictor variables per country (description, categorization, and source in S2). Vector and pathogen hazards were based on habitat suitability projections for phlebotomine sand flies (at a spatial resolution of 30 arcsecond, mean of 1980-2010) and *Leishmania* prevalence in sand flies and vertebrate hosts (at NUTS 2-3 level, (multi-)annually between 2002-2024). The abundance of an important domestic reservoir was based on the registered number of cats and dogs, but these data were only available for Portugal (at NUTS3 level, annually in 2010-2024) and Italy (at NUTS2 level, snapshot in April 2025). We used 78 climatic variables (at spatial resolution of 0·5 degree, mean of 1979-2018, and mean of 2040-2060, 2060-2080 and 2080-2100 from the general circulation model ACCESS1-0 for the Representative Concentration Pathway (RCP) 4·5 and 8·5) from the Climatic Data Store (https://cds.climate.copernicus.eu). We also used 14 land-use variables (at spatial resolution of 0·25 degree and annually from 850 to 2019) from NASA Earthdata (https://search.earthdata.nasa.gov/search) and 22 biodiversity indicators (at spatial resolution of 1 degree, snapshots in 1900, 2015, 2050 and additional for birds 1900-2015 decadal) from The Group on Earth Observations Biodiversity Observation Network (GEO BON, https://geobon.org/). The land-use variables distinguished five types of crop land, managed pasture, rangeland, forested and non-forested primary land, potentially forested and non-forested secondary land, the age of secondary land, the biomass of secondary land, and urban land (S2). The biodiversity indices included the absolute and relative changes between snapshots in species richness of birds, mammals, and vertebrates overall and the intactness of vertebrate communities, as vertebrates can serve as reservoir. (2) Socioeconomic variables included livestock farming intensity, human population size, demography, education level, housing conditions, inequality, disposable income, poverty and social exclusion, unemployment, occupation, and hospital capacity. Human immunosuppressive conditions consisted of HIV incidence and mortality rate, and various cancer incidences and mortality rates. These data, which were at NUTS 2 or 3 level and included both snapshots and multi-annual runs between 1995 and 2023, mainly came from Eurostat (https://ec.europa.eu/eurostat/web/main/home). We collected human population density (at spatial resolution of 30 arcseconds and temporal resolution of 5-year intervals between 2000 and 2020) from NASA’s Socioeconomic Data and Applications Center (SEDAC, https://earthdata.nasa.gov/centers/sedac-daac) and human footprint index (at spatial resolution of 1km and temporal resolution of 1 year in 2000-2022). Age- and sex-stratified population data in Spain (at spatial resolution of NUTS3 and temporal resolution of 1 year in 1997-2015) were collected from the National Institute of Statistics (https://ine.es/).

### 2.3 Modelling process

The modelling procedure consisted of three steps: First, in the preprocessing phase, leishmaniasis incidences and the environmental variables were mapped to the NUTS 2024 structure. (9) Annual incidence data (i.e., Spanish and Italian datasets) were summed to cumulative incidences to remove temporal fluctuation. When reported incidences included sex and age (group) attributes (i.e., Spanish dataset), we computed SIRs as outlined in section 2.1. Afterwards, we extracted the values of predictor variables for those NUTS with reports (including zero cases). We matched the temporal extents of the predictor variables with the response variables (S2). Predictor variables with higher spatial granularity than NUTS3 (i.e., sand fly habitat suitability, climate, land use, biodiversity, human population density, human footprint index) were aggregated to the level of NUTS2 for Italy and NUTS3 for other countries, where the values of the fully and partially enclosed grid cells were averaged. Other predictor variables were matched to the NUTS level of the response variables (Table 1). All predictor variables were standardized.

Second, in the processing phase, we fitted models and computed model statistics and effect sizes. For Spain, we correlated the number of hospital discharges of patients with HIV-leishmaniasis comorbidity with the number of hospital discharges of patients with leishmaniasis regardless of other diagnosis. We also tested the impact of sex and age group on leishmaniasis incidence using beta-binomial regression and then Tukey-adjusted z-tests for pairwise comparisons. For each study country, we fitted regression models. We started with fitting two null models that only contained an intercept and spatial effect: one null model assumed neighbor structure based on contiguity with conditional autoregression, (13) and the other null model assumed a distance-based effect captured by stochastic partial differential equations (SPDE) (S3). (14) The first would be appropriate, for example, for policy convergence in adjacent jurisdictions; (15) the second, when disease transmission is a function of distance because of vector dispersal distance. (2) Then, using each predictor variable one at a time (except for variables with sample size ≤ 15, which we deemed too few to reliably detect an effect), six models were fitted: a generalized linear model (GLM), a GLM with neighbor structure spatial effect, a GLM with distance-based SPDE spatial effect, a generalized additive model (GAM), a GAM with neighbor structure spatial effect, and a GAM with distance-based SPDE spatial effect (S3). The GLM distributions depended on the dataset (Table 1). Watanabe-Akaike Information Criterion (WAIC) was computed for all fitted models. In addition, the mean and the 0·025 and 0·975 quantiles of the coefficient estimates were computed for the three linear models (i.e., GLMs) for each variable.

For France, where many NUTS3 regions reported zero cases, a zero-altered Poisson (ZAP) model (S3) was fitted. A ZAP model contained a Bernoulli model to classify presence / absence of leishmaniasis and a zero-truncated Poisson (ZTP) model to regress leishmaniasis incidence in leishmaniasis-present regions onto the predictor variables. The sample size requirement for the ZTP models was relaxed to 14, since only 14 French NUTS3-regions reported leishmaniasis. We fitted one-variable Bernoulli models and single-variable ZTP models and conducted variable selection. For Bernoulli models, a variable was selected if the WAIC of one of the six models was lower than the WAIC values of both null models by at least 2 (16). For the ZTP models, the same criterium was applied and it was only met by four variables: three education levels and one climatic variable. To limit the number of ZAP models, we selected the education variable with the lowest WAIC (which also had the largest effect size) and the climatic variable. We fitted ZAP models by combining one selected (of the 254) variable in Bernoulli model and one (of the two) selected variable in ZTP, for all possible combinations.

For all countries, after fitting the single-variable models, we selected one variable per category (S2) with the lowest WAIC and that must be lower than the WAICs of the null models by 2. (16) Then, each variable from those selected was paired in turn with every variable from the full set to fit six models (i.e., the three GLMs and the three GAMs) containing both variables and their interaction, except when VIF was ≥ 5, (17) sample size was ≤ 45 (15 observations per variable / interaction as above), or both were true. For France, we fitted ZAP models with two-variable combinations in the Bernoulli part and a single variable in the ZTP part. When the sample size for a selected variable was ≤ 45, a substitute (the most correlated variable with sample size >45 and with a correlation coefficient ≥ 0·8 or ≤ −0·8) was used. WAIC was computed per model for each variable combination.

Third, in the post-processing phase, we identified risk factors per country based on WAIC (i.e., models with WAIC values that were less than the WAIC values of both null models by at least 2) and the coefficient estimates of the linear models. The model with the lowest WAIC and good fit for all Spanish continental NUTS3 regions was used for geographic mapping of relative risk (i.e., risk excluding demographic structure) in Spain (i.e., excluding Balearic Islands due to lack of predictor variables). The best fitting model (with the lowest WAIC) for continental France (i.e., excluding Corsica due to lack of predictor variables) contained a climatic variable, which enabled future projection. Future projection was not possible elsewhere, as the best fitting models for the other countries only contained variables that currently do not have future projections (i.e., socioeconomic variables; biodiversity variables; pathogen-, vector-, and reservoir-related variables). Hence, we did not generate maps for Greece, Italy, and Portugal.

## 3. Results

### 3.1 Standardized incidence ratio in Spain

Between 1997 and 2015, Madrid reported the most leishmaniasis cases in Spain (Fig. 1a). After adjusting for population and sex- and age-structure, the highest relative risk for leishmaniasis was in Jaén with SIR = 6·26 (Fig. 1b), followed by Córdoba (SIR = 2·65) and Toledo (SIR = 2·10). An additional 10 NUTS3 regions had SIR > 1 including Madrid (SIR = 1·74) and eastern coastal regions. The best fitting model (Fig. 1c) contained human footprint index and species richness change of all vertebrates between 2015 and 1900. Interactive maps are in S4.

**Figure 1.**
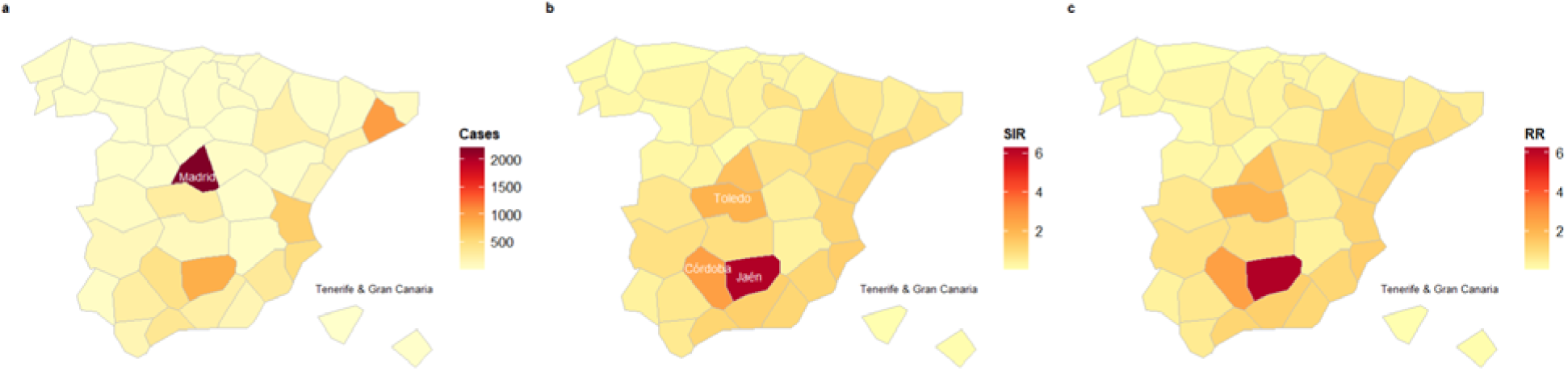
Reported leishmaniasis cases (a), standardized incidence ratio (SIR, b), and fitted relative risk (RR, c) in Spain (excluding Balearic Islands) between 1997-2015.

### 3.2 Risk factor analysis

The Spanish datasets contained information on HIV status, sex and age, allowing for analysis of HIV-leishmaniasis comorbidity and comparison of leishmaniasis risks among populations with different sex-age structures. HIV-leishmaniasis comorbidity made up two thirds of hospital discharges of adult leishmaniasis patients (i.e., ≥ 20 years old) and 58·8% of all ages. Considering all NUTS3 regions, sex and age groups, the number of hospital discharges of patients with HIV-leishmaniasis comorbidity were highly correlated with that of leishmaniasis overall (Pearson’s correlation, ρ = 0·83, df = 518, p-value < 0·001). In patients without HIV, leishmaniasis remained a pediatric disease (Fig. 2). Female and male children under the age of five had two to six times higher risk for leishmaniasis compared to other age groups (Fig. 2)

**Figure 2.**
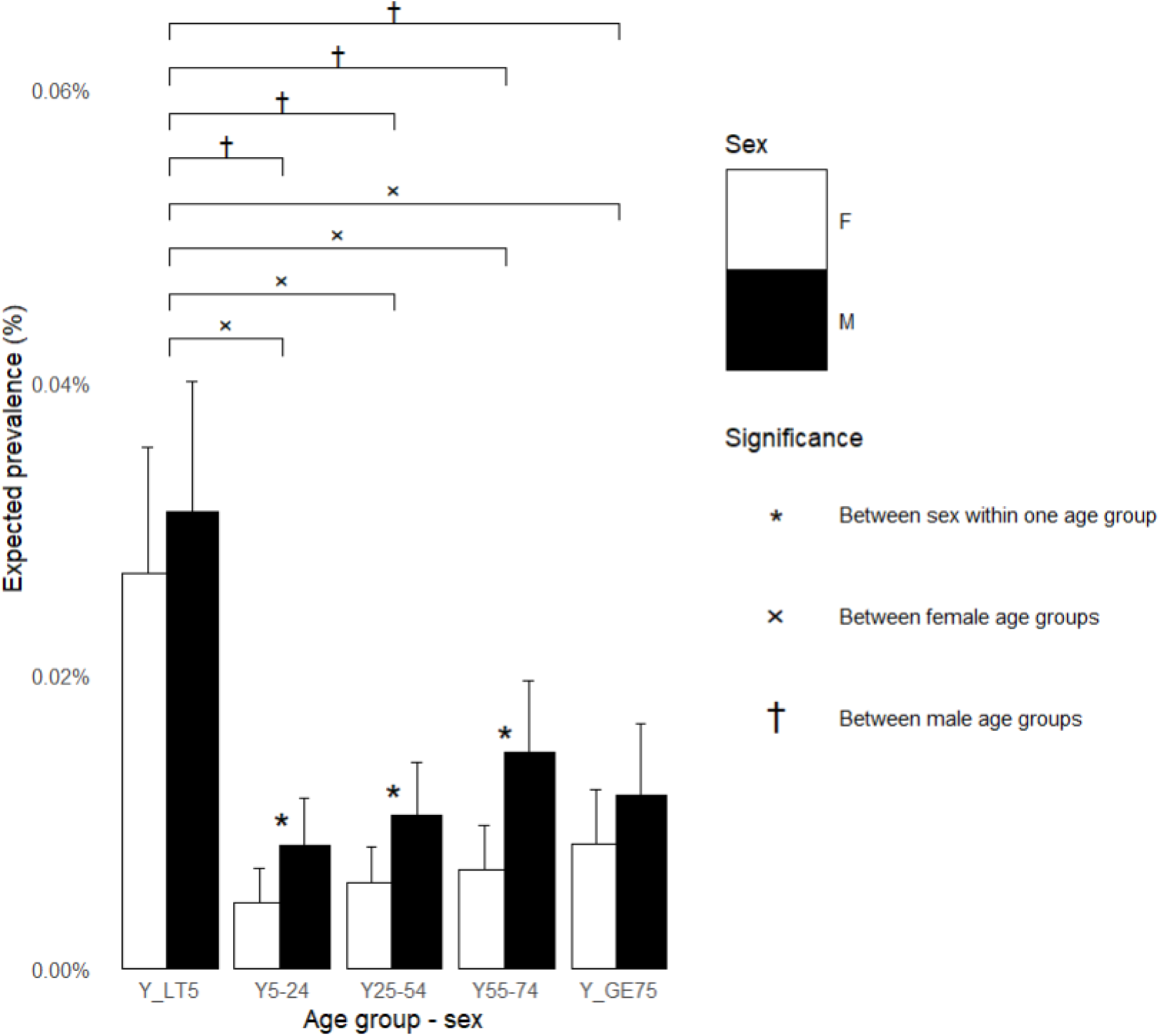
Expected leishmaniasis prevalence (%) excluding HIV comorbidity per age group and sex in Spain. Error bars reflect 95% confidence interval; stars indicate statistical significance (α = 0·05). Y_LT5 = less than 5 years old; Y5-24 = 5 to 24 years old; Y25-54 = 25 to 54 years old; Y55-74 = 55 to 74 years old; and Y_GE75 = 75 years old or higher.

Risk factors varied across countries (Fig 3). In Greece, the most influential (i.e., with the largest effect size) risk factors were socioeconomic: occupation in sector agriculture, forestry, and fishery followed by housing type (i.e., one dwelling conventional residential building) (Fig. 3a). Occupation was also a risk factor in France, Spain, and Portugal, although the sector that was associated with leishmaniasis risk or protection varied per country (Fig. 3c-e). Besides in Spain (Fig. 2), being immunocompromised was found to be an effective risk factor in Italy (Fig. 3b). In Italy, leishmaniasis was positively correlated with the 3-year average death rates of various cancer types, except for malignant neoplasm of upper gastrointestinal tract, brain and central nervous system, and thyroid gland, all three of which showed negative correlations. Climate, the most influential factor in France, was only of secondary importance in Greece and Italy (Fig. 3c). In these countries, warmth and dryness were positively associated with leishmaniasis incidences. In the Iberian Peninsula, climate did not provide more information about leishmaniasis than spatial autocorrelation. *Leishmania* prevalence in dogs was a risk factor in all countries where these data were available. Dog and cat population sizes were risk factors in Portugal. The change of species richness of forest birds between 2010 and 2000 negatively related to leishmaniasis in France and Greece (Fig. 3a, 3c).

**Figure 3.**
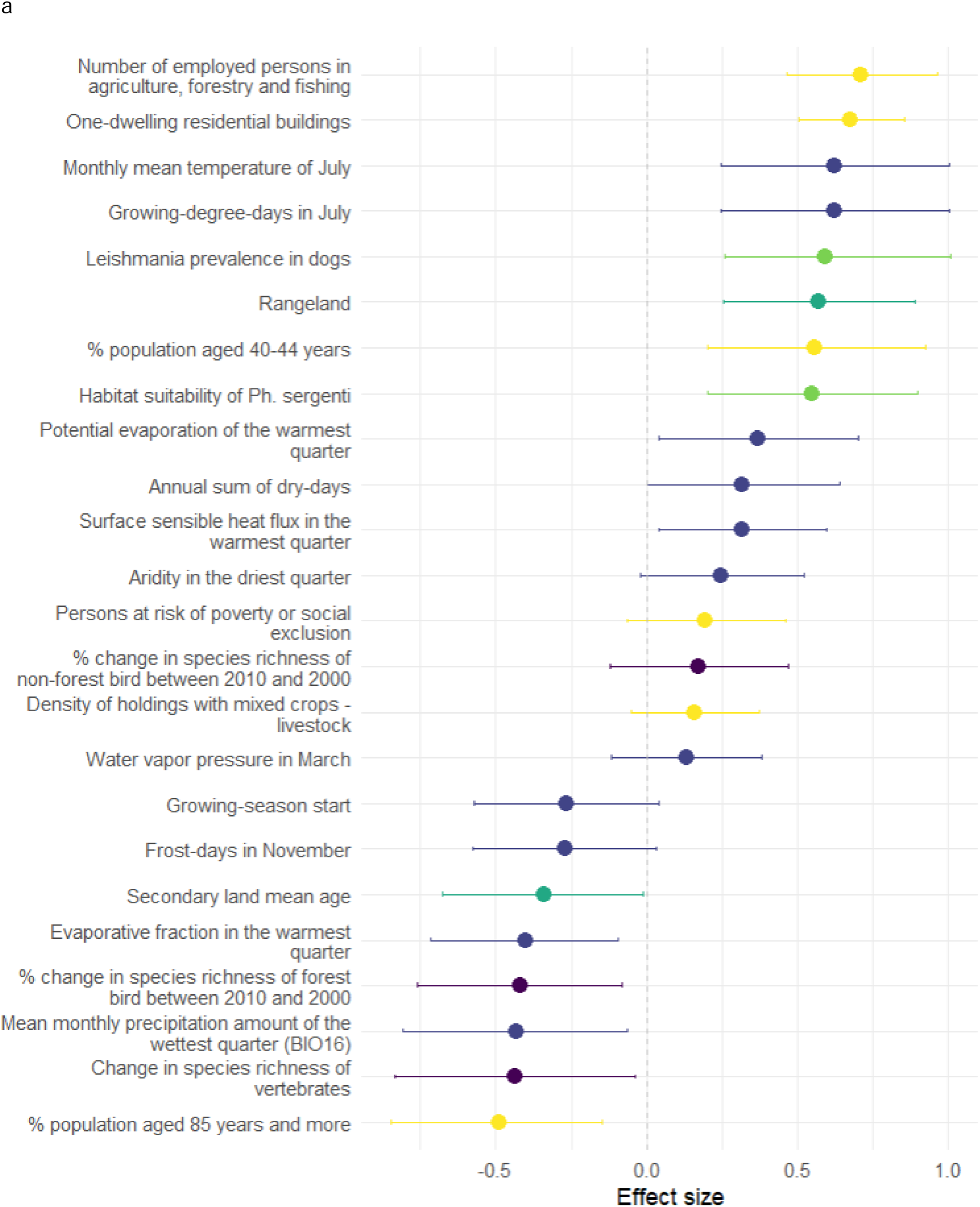
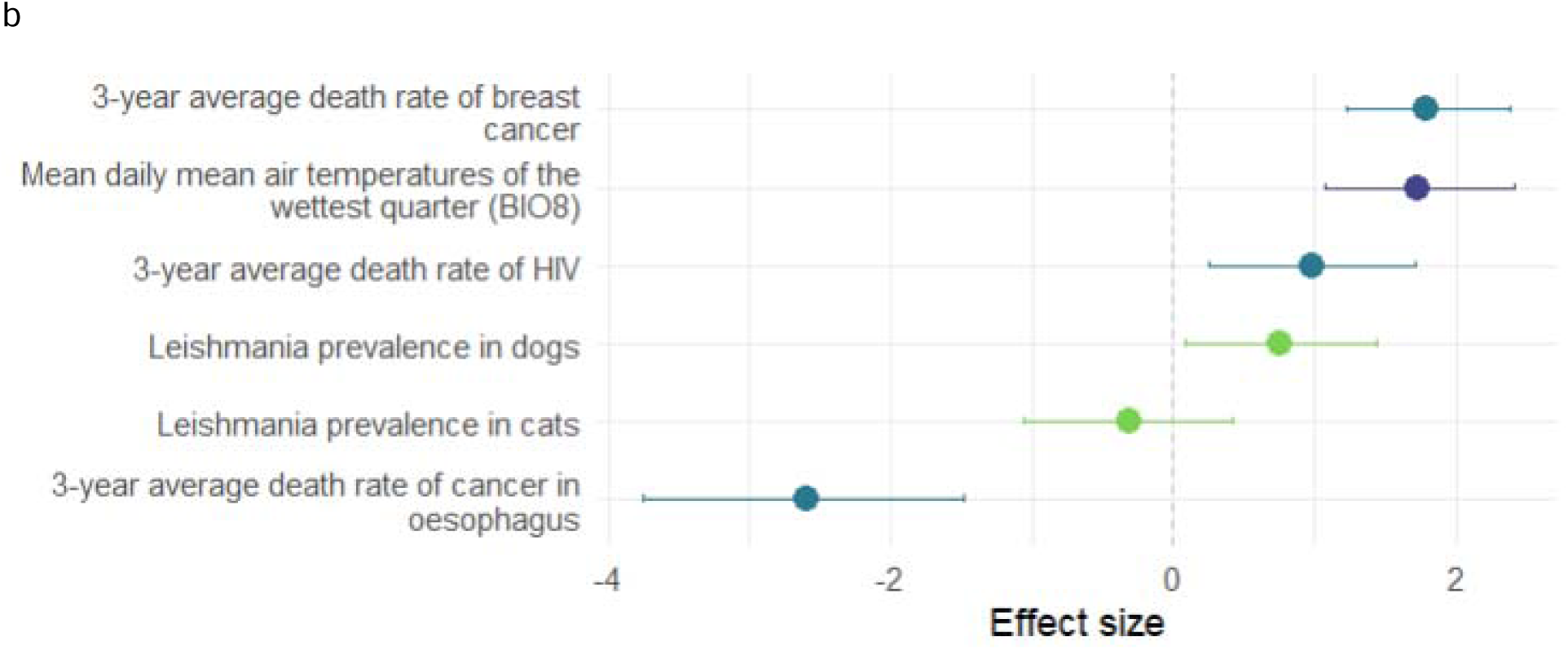
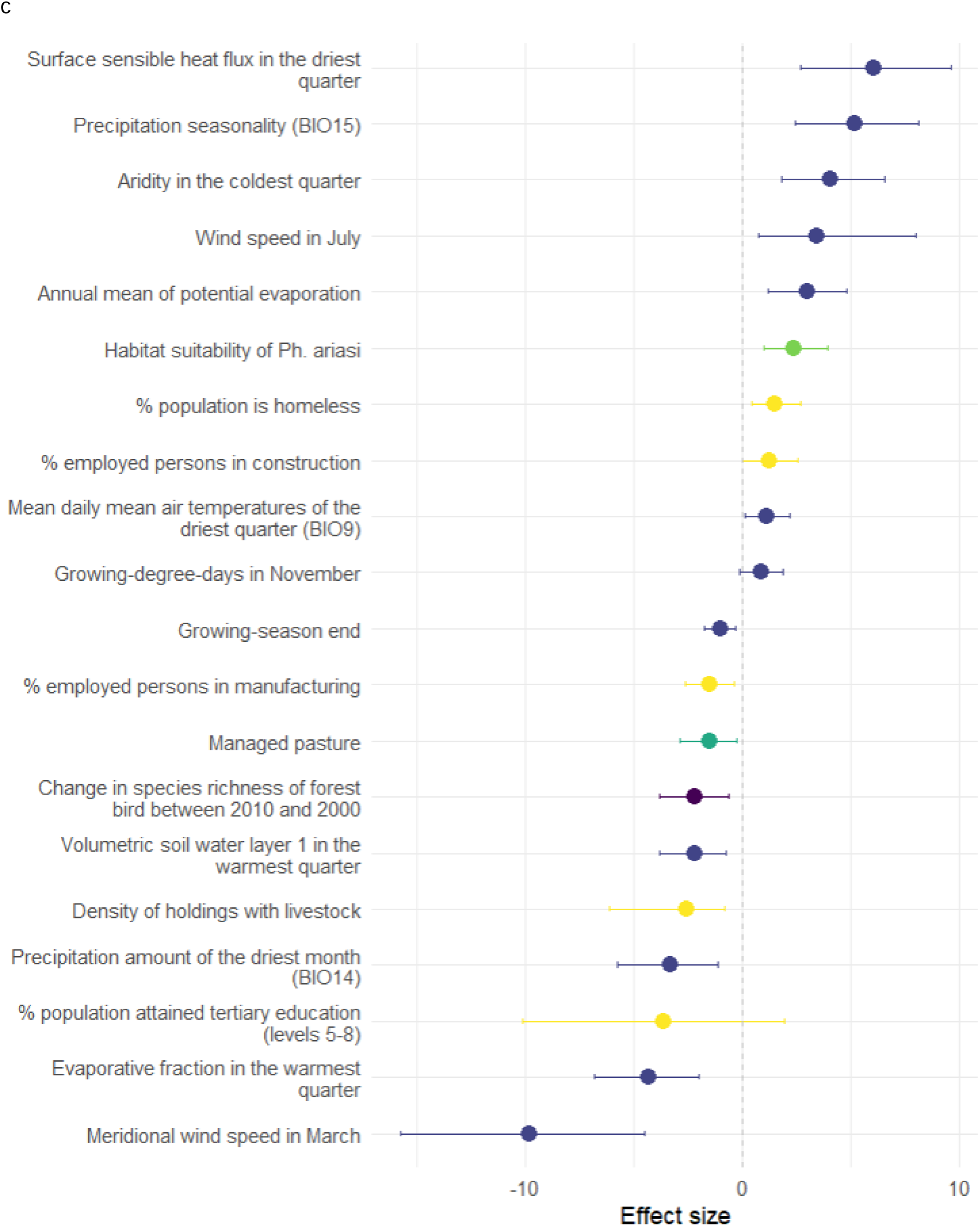
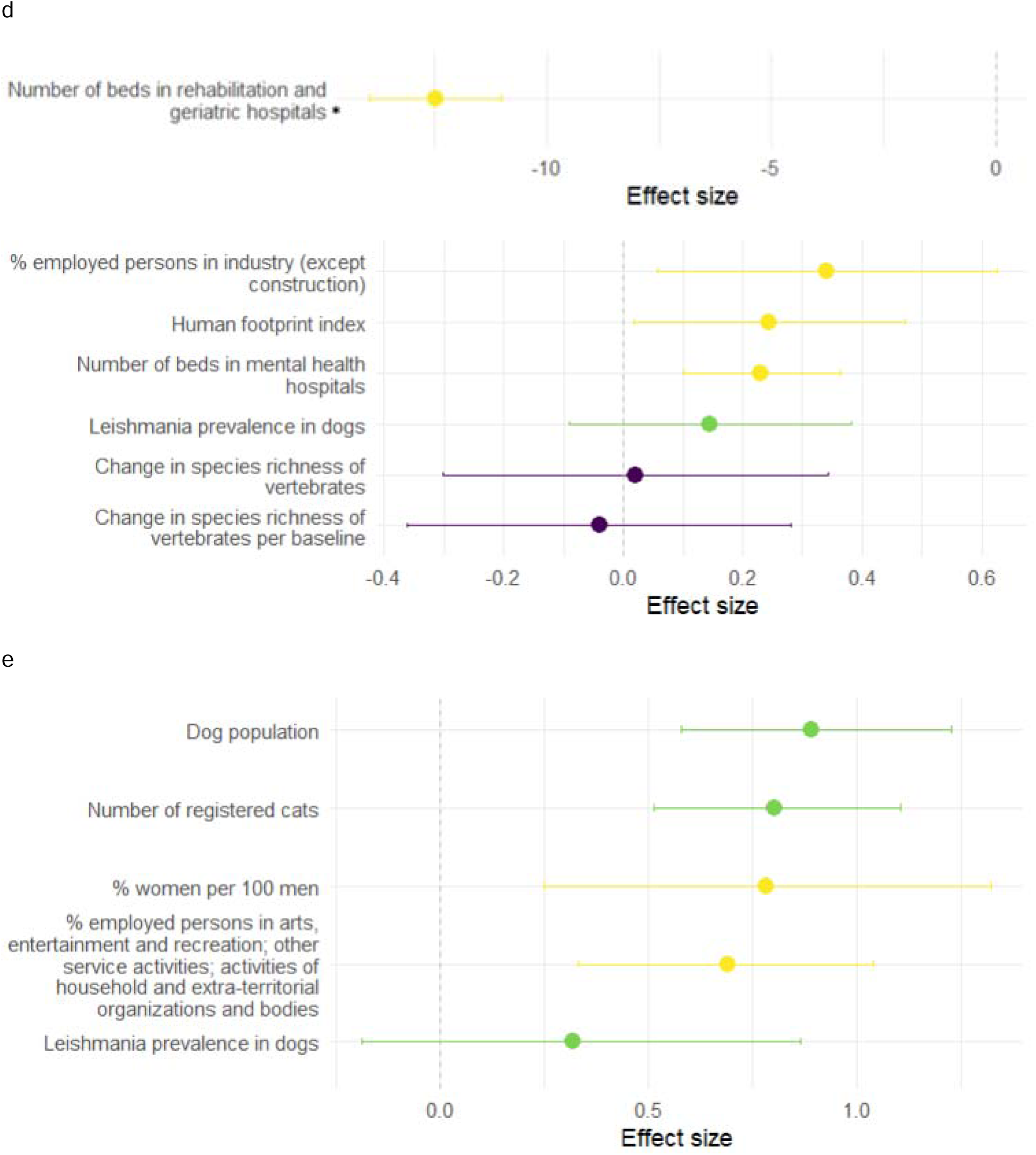
Selected risk factors of leishmaniasis in Greece (a), Italy (b), France (c), Spain (d), and Portugal () showing mean estimates and 95% credibility intervals. If variables within a category (S2) positively or negatively correlated with leishmaniasis, the variable with the largest effect size is shown. If within a variable category both positive and negative links to leishmaniasis were found, the variables with the largest positive and the largest negative effects are shown. Colors distinguish variable groups (S1): dark purple = biodiversity, blue = climate, grey blue = human immunosuppressive condition, teal = land use, green = pathogen-, vector-, and reservoir-related, yellow = socioeconomic. The same sets of biodiversity, climatic, and land-use variables were tested in all countries, whereas human immunosuppressive condition; pathogen-, vector-, or reservoir-related variables; and socioeconomic variables varied among countries and were tested as available (S2). HIV = human immunodeficiency virus. *Ph*. = *Phlebotomus.* * This risk factor is plotted on its own scale for greater clarity.

### 3.3 Prediction in France

Future prediction of leishmaniasis incidence was limited to France by the availability of future projections of predictor variables. In France, when controlling for all other conditions, climate change was not expected to drive emergence of leishmaniasis in non-endemic regions within this century (Fig. 4).

**Figure 4.**
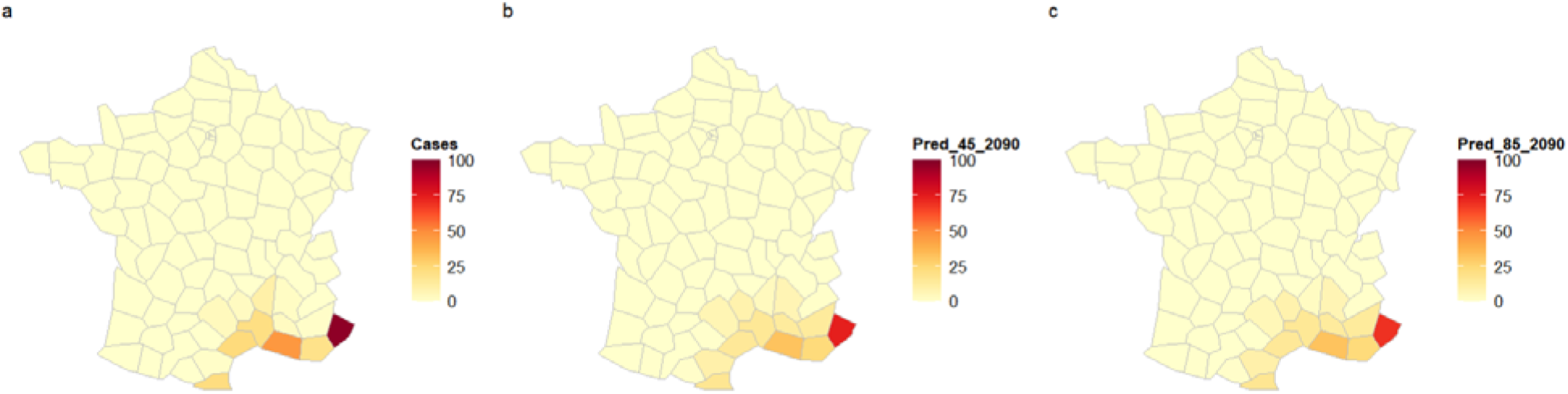
Reported leishmaniasis incidence in France (excluding Corsica) in the period 1999-2012 (a) and predicted incidence for the period 2081-2100 under the Representative Concentration Pathway (RCP) 4·5 (b) and RCP 8·5 (c)

## 4. Discussion

Leishmaniasis is an endemic disease in the European Mediterranean region with high risks of outbreaks and geographical spread (2), although the incidence and disease burdens are relatively low compared to the Indian subcontinent, Brazil, East Africa, North Africa, and the Middle East. (1) Possibly due to the relatively low burden, risk factors of leishmaniasis in Europe have been understudied. (3) Here, we tested the importance of a large variety of potential risk factors of leishmaniasis in five southern European countries. We found that socioeconomic factors and human immunosuppressive conditions affected leishmaniasis incidence the most in European Mediterranean countries where leishmaniasis is endemic, although the specific variables varied among the studied countries (partly due to data availability), as did their impact. For the socioeconomic factors for example, in Greece people working in sector agriculture, forestry and fishing had higher chance to be exposed to hazard (Fig. 3). In France, workers in construction had higher and in manufacturing lower risk for leishmaniasis, possibly due to differences in outdoor and indoor activities. In contrast, in Spain the highest risk occupation sector was industry (including manufacturing but excluding construction). The Spanish results differ from those of other counties in part because the response variable in Spain (i.e., SIR) was adjusted for demographic structure. In addition, the high-risk sectors in Portugal (i.e., arts, entertainment and recreation, other service activities, activities of household, and extra-territorial organizations and bodies) suggest that people in smaller economic sectors can as well benefit from targeted public health efforts. The identified socioeconomic risk factors in our study largely differ from those in the above-mentioned regions outside of Europe. These differences are probably partly due to study designs and methodology. Studies at much smaller spatial scales (e.g., village or city level) can include variables like sanitary services or ownership of (infected) dogs, which are not available at the country level. However, there are also real global differences that complicate comparisons. Unlike in Europe, regions that lack basic civil facilities such as sewage services, water supply, and garbage collection and where houses made of straw and mud, provide ideal habitat for sand flies. (3) Furthermore, being malnourished and immunocompromised can increase the vulnerability of susceptible populations. (3) Those factors, which are often associated with some combination of low education level, poverty, and migrant status, increase the risk of leishmaniasis. (3) Our study identified certain housing conditions, such as one-dwelling residential buildings in Greece and being homeless in France, as risk factors (Fig. 3A, 3C). Although the reasons for the high-risk profiles of these two conditions require further study, it is plausible that one-dwelling residential buildings are likely to be found in peri-urban and rural areas, guarded by dogs living outdoors and exposed to sand flies, raising the risk for leishmaniasis. Meanwhile, people experiencing homelessness often live outdoors and may also be malnourished, translating to both greater exposure and vulnerability for leishmaniasis. Interestingly, we found no correlation between poverty (i.e., at-risk-of-poverty rate, persons at risk of poverty or social exclusion, S2) and leishmaniasis incidence.

Infections and other health conditions that suppress the immune system are thought to partly underlie a demographic shift of leishmaniasis patients in Europe. In recent decades in this region, leishmaniasis has gone from being a pediatric disease to a disease mostly affecting immunocompromised adults. (18) Indeed, we found that adult males in Spain (i.e., 25-54 years old) and in Greece (i.e., 40-44 years old) had the highest risks. In Spain, the number of hospital discharges of patients with HIV-leishmaniasis comorbidity accounted for two-thirds of the hospital discharges of adult leishmaniasis patients. Our results align with earlier reports that document links between HIV and leishmaniasis. (18) Furthermore, a nuanced picture emerged from Italy, where leishmaniasis was positively associated with the mortality rates of some types of cancer (e.g., breast cancer, lymphoma, leukemia) and negatively associated with other types (e.g., malignant neoplasm in esophagus, brain and central nervous system). The positive associations are in line with clinical observations of coincidence of these diseases and may relate to heightened susceptibility associated with immunosuppression due to cancer therapy. (19) A similar positive correlation was also found between malaria incidence and all cancer mortality in the U.S.. (20) The negative associations might be linked via physiological or biochemical mechanisms. (21) Similar antagonistic relationships were also found between cancer and other protozoan parasites. (22) More generally, the elicitation of immune responses to infection with *Leishmania* species or tumor formation and growth might offer protection against the other disease. (23) Importantly, because they came from highly aggregated data (i.e., NUTS level-2) and thus a small sample size (N=19 NUTS level-2 administrative units in Italy), our results should be seen only as a heuristic and not as an answer to the complex question of cancer-leishmaniasis co-morbidity.

A previous study predicted that climatic conditions would become more suitable for *Leishmania infantum* in current endemic regions in Europe and that suitable areas will expand northwards. (4) However, our results and predictions for France, the only country in our study that climate delineated endemic and non-endemic regions, contradicted that prediction: our analyses suggested no expansion of leishmaniasis into currently non-endemic regions by the end of this century under two RCPs. In our other studied countries, climate had less effect than socioeconomic conditions. While the earlier study focused on climate suitability, (4) in the current study we evaluated variables related to climate, land use, biodiversity, socioeconomic conditions, and pathogens, vectors, and reservoirs. By employing this approach, we were able to assess the relative importance of climate, and other variables, to leishmaniasis risk. In Greece, Italy, and France, but not in Spain or Portugal, we found that warmth and dryness were positively associated with leishmaniasis incidence. The positive association of warmth on leishmaniasis incidence could work via improved vector habitat suitability and increased vector activity, which are temperature-related and also found in other vector-borne diseases (e.g., tick-borne borreliosis, West Nile Virus infection). (24) While previous studies have highlighted the importance of moisture to sand flies during immature stages (2) and in characterizing suitable sand fly habitat, (25) our results indicated a role for dryness on a larger scale. This finding aligns with field observations that sand flies in Europe occupy warm and dry macroenvironments and adult abundance is often associated with low relative humidity. e.g., 26 Dryness was positively correlated with hazard risk, and this relationship is seemingly relevant both during summer when adult sand flies are active and during winter when larva enter diapause. In addition, the role of warmth and dryness in facilitating both human outdoor activity and vector activity (24) should not be overlooked, since together these types of activity compound the chances of exposure. In the Iberian Peninsula, our model did not detect a relationship between climate and leishmaniasis cases. In this case, the absence of a relationship with climate might be a consequence of the spatial term in the model accounting for patterns in both climate and reported cases; if so, climate would not provide extra information above spatial autocorrelation (i.e., closer things are more related) for understanding the disease risk. Indeed, non-spatial models with one or two climatic variables (the number of variables is limited by the sample size) had poorer model performance (i.e., higher WAIC) than null models (that were spatial models), indicating spatial structure explains and predicts disease risk better than climatic variables in the Iberian Peninsula.

Compared to vector hazard, pathogen hazard and reservoir abundance were relatively more important risk factors for leishmaniasis. In Portugal, Spain, and Italy, leishmaniasis incidence was not related to the habitat suitability of sand fly vectors, the proxy for vector hazard. Furthermore, in France, leishmaniasis was only reported in southeastern regions (8) despite *Ph. perniciosus*, an important vector of visceral leishmaniasis, also occurring in middle and northern France, (27) albeit at a low density. Climate delineated regions in France with and without leishmaniasis, possibly by regulating pathogen presence. However, pathogen hazard could not be assessed in France due to lack of data. In the other four countries, pathogen hazard was identified a risk factor of leishmaniasis. Similarly, reservoir abundance (e.g., dog and cat population) was found to be an effective risk factor in Portugal. Our findings highlight the need for surveillance of *Leishmania* species and assessment of reservoir species in addition to vector surveillance. These data should be made widely accessible for the purpose of disease prevention and preparedness.

The relationship between biodiversity variables with leishmaniasis risk was mixed (Fig. 3). In general, in France and Greece where biodiversity variables were identified as risk factors, the negative relationships were stronger than the positive ones. A decrease in forest bird diversity was associated with an increase in leishmaniasis risk in both countries; changes in total vertebrate diversity (i.e., including birds) was also inversely related to leishmaniasis risk in Greece. These results suggest that leishmaniasis risk could be governed by host community structure or other mechanisms underlying biodiversity-disease relationships, like the dilution effect. (28) A previous study on Amazonian leishmaniasis in French Guiana demonstrated that higher mammal diversity was associated with lower *Leishmania* prevalence in vectors but with higher sand fly density, suggesting biodiversity simultaneously reduced pathogen hazard and increased vector hazard. (29) The role of biodiversity on other vector-borne diseases, particularly tick-borne borreliosis, has been intensely studied and debated in both human-dominated and more natural environments. (28) A synthesis of these studies suggests that although biodiversity-disease relationships are nonlinear and scale-dependent, biodiversity regulates disease outbreaks at scales most relevant for public health (i.e., < 100 km^2^, roughly the size of a small city). (28) Our findings support the rationale of One Health approach: By integrating wildlife and their ecology, leishmaniasis research can gain insights into topics such as sylvatic cycles, pathogen distribution, reservoir competence, and, more generally, the regulatory roles of biodiversity on leishmaniasis.

The diverse nature of the risk factors included in our study gives rise to an important caveat: variables relate to risk in different ways and to different degrees. For example, an increased population with “risky” occupations means more people (i.e., employees in these sectors) have a higher chance to encounter a hazard, whereas a rise in temperature of the same magnitude can affect the whole susceptible population and even generate new hazards. Consequently, comparing effect sizes alone may fall short. An important next step towards understanding the relative importance of risk factors could be scenario-based modelling using potential impact fraction (PIF).(30) In the scenarios, the modifiability of the risk factors could be integrated to simulate realistic, partial reductions in risk, offering a more tangible metric for policy planning.

Overall, our analyses suggest that the risk factors for leishmaniasis in the endemic regions in Europe are primarily socioeconomic and human immunosuppressive conditions. To date, no large-scale analysis has been carried out, particularly one that includes diverse environmental variables beyond only climate variables, in the areas with high leishmaniasis burden, such as the Indian subcontinent, Brazil, East Africa, North Africa, and the Middle East. (3) Despite differences in socioeconomic conditions and spatial scales, our analyses agree with analyses of data from regions with high disease burdens: socioeconomic variables are major risk factors for leishmaniasis. We suggest that in southern Europe adaptive control efforts should be context specific and target vulnerable groups (e.g., information and protective measures for immunocompromised population, specific age groups and occupations) because risk factors for leishmaniasis varied from country to country and among populations. In addition, broadly impactful measures such as poverty alleviation and climate change mitigation could also help reduce leishmaniasis outbreaks and geographical spread. Finally, surveillance of *Leishmania* in human and non-human vertebrates and the accessibility of these data will help to better understand this parasite and protect people from this disease.

## Data Availability

All data produced in the present work are contained in the manuscript.

## Acknowledgment

The transfer and use of the Spanish datasets were administrated with a memorandum of understanding (MoU) supplementing the grant agreement of EU-Horizon project “Climate Monitoring and Decision Support Framework for Sand Fly-borne Diseases Detection and Mitigation with Cost-benefit and Climate-policy Measures” (CLIMOS, https://climos-project.eu/). We thank Daniela Nunes for the administration of MoU and data transfer agreements.

This study used data provided by Portuguese Companion Animal Information System (SIAC).

## Author Contribution

DW conceived the idea, designed the study, constructed the dataset, conducted the analysis and drafted the manuscript. KDM, ARH and FL conceived the idea, designed the study and commented on the manuscript. CM acquired closed datasets and commented on the manuscript. EB provided closed dataset and commented on the manuscript, JR, EVS, AX, GC, FV and FB provided closed datasets. All authors read and approved the manuscript.

## Data sharing statements

Data sources were disclosed in the text and S2.

## Supporting information captions

S1. R functions and packages

S2. Predictor variables

S3. Model formulation

S4. Interactive map of relative risk of human leishmaniasis in Spain (excluding Balearic Islands) in 1997-2015.

