## Supporting information S1-S3 for "Socioeconomic risk factors and human immunosuppressive conditions are major drivers of human leishmaniasis in southern Europe"

### S1. R Functions and Packages

| Variable | Description | Function | Package |
|---|---|---|---|
| SIR | Computing the expected cases of leishmaniasis in Spain, given reference population (i.e., national population) per sex-age group, and the population structure per region (i.e., on NUTS3 level.) | expected() | SpatialEpi (1) |
| sand fly habitat suitability, climate, land use, biodiversity, human population density, human footprint index | Aggregating predictor variables with high spatial granularity to NUTS 2 or 3 level. | extract() | terra (2) |
| All predictor variables except above | Matching predictor variables to response variables in space | left_join() | dplyr (3) |
| HIV | Testing the correlation between HIV-leishmaniasis hospital discharges and overall leishmaniasis hosptial discharges in Spain | cor.test() | stats (4) |
| sex, age group | Regression of sex and age-agroup on leishmaniasis hosptital discharges in Spain | glmmTMB() | glmmTMB (5) |
| sex, age group | Pairwise comparison of expected leishmaniasis prevalence amoung sex and age-groups in Spain | emmeans(), pairs() | emmeans (6) |
| All predictor variables seperately and pairwise combination with selected variables | Baysien regression of predictor variable(s) on reported leishmaniasis incidence in Portugal, France, Italy and Greece | | INLA (7–9) |
| Selected predictor variables | Plotting effect sizes | | ggplot2 (10)<br>viridis(11)<br>forcats(12) |
| Reported and fitted leishmaniasis risk | Static mapping of Spain and France | | ggplot2 (10)<br>sf(13)<br>RColorBrewer(14) |
| Reported and fitted leishmaniasis risk | Interactive mapping of Spain | | sf(13)<br>mapview(15)<br>RColorBrewer(14)<br>leafpop(16)<br>leafsync(17) |

**S2. Predictor Variables**

| Group | Dataset | Category | Variable name | Unit | Temporal span | Temporal resolution | Spatial resolution | Spain 1997-2015 | France 1999-2012 | Italy 2011-16 | Greece 2009-18 | Portugal 2010-20 |
|---|---|---|---|---|---|---|---|---|---|---|---|---|
| pathogen-, vector-, and reservoir-related | SF suitable habitat(18) | SF habitat suitability | habitat suitability | probability | m(1980-2010) | snapshot | 30 arcsec | near-present | near-present | near-present | near-present | near-present |
| | | | binary habitat suitability | | | | | near-present | near-present | near-present | near-present | near-present |
| | *Leishmania* prevalence in SF(19) | *Leishmania* prevalence in SF | *Leishmania* prevalence in SF | % | 2002-2022 | multi-annual | NUTS2-3 | 2002-2018 | | 2008-2022 | 2009-2020 | 2007-2017 |
| | CanL(19) | *Leishmania* prevalence in dogs | *Leishmania* prevalence in dogs | % | 2005-2022 | multi-annual | NUTS2-3 | NA | | 2017-2019 | 2005-2017 | 2022 |
| | CanL in Italy (unpublished data) | | *Leishmania* prevalence in dogs in Italy | % | 2010-2023 | annually | NUTS2 | | | 2011-2016 | | |
| | CanL in Portugal (unpublish-ed data) | | *Leishmania* prevalence in dogs in Portugal | % | 2011-2024 | annually | Municipality, NUTS3 | | | | | m(2011-20) |
| | FelL(19) | *Leishmania* prevalence in cats | *Leishmania* prevalence in cats | % | 2009-2019 | multi-annual | NUTS2-3 | NA | | 2017-2019 | 2009-2015 | |
| | Dog population in Portugal (20) | Pet population | dog population in Portugal | number | 2020 | snapshot | NUTS3 | | | | | 2020 |
| | Registered pets in Italy(21) | | number of registered cats in Italy | number | 6-4-2025 | snapshot | NUTS2 | | | 2025 | | |
| | Registered pets in Portugal (unpublished data) | | number of registered dogs and cats in Portugal | number | 2010-2023 | annually | NUTS3 | | | | | cat 2020; dog m(2011-20) |

| | | | | | | | | | | | | |
|---|---|---|---|---|---|---|---|---|---|---|---|---|
| | *Leishmania* prevalence in livestock (19) | *Leishmania* prevalence in livestock | *Leishmania* prevalence in livestock | % | 2007-2016 | multi-annual | NUTS2-3 | | | | 2007-2016 | |
| | *Leishmania* prevalence in wildlife(19) | *Leishmania* prevalence in wildlife | *Leishmania* prevalence in wildlife | % | 2007-2016 | multi-annual | NUTS2-3 | NA | | NA | 2007-2016 | NA |
| Socio-economic | Agriculture (22) | Agriculture | Density of holdings with livestock | number / km2 | 2000-2007 | bi-annually | NUTS 3 | mean | mean | latest | latest | latest |
| | | | Density of holdings with bovine animals | number / km2 | | | | | | | | |
| | | | Density of bovine animals | number / km2 | | | | | | | | |
| | | | Density of holdings with dairy cows | number / km2 | | | | | | | | |
| | | | Density of dairy cows | number / km2 | | | | | | | | |
| | | | Density of holdings with other cows | number / km2 | | | | | | | | |
| | | | Density of other cows | number / km2 | | | | | | | | |
| | | | Density of holdings with sheep | number / km2 | | | | | | | | |
| | | | Density of sheep | number / km2 | | | | | | | | |
| | | | Density of holdings with goats | number / km2 | | | | | | | | |
| | | | Density of goats | number / km2 | | | | | | | | |

| | | | | | | | | | | | |
|---|---|---|---|---|---|---|---|---|---|---|---|
| | | Density of holdings with pigs | number / km2 | | | | | | | | |
| | | Density of pigs | number / km2 | | | | | | | | |
| | | Density of holdings with poultry | number / km2 | | | | | | | | |
| | | Density of poultry | number / km2 | | | | | | | | |
| | | Density of holdings with mixed livestock | number / km2 | | | | | | | | |
| | | Density of holdings with mixed crops - livestock | number / km2 | | | | | | | | |
| | | Portion of agriculture area of holdings with mixed livestock | agriculture area / km2 | | | | | | | | |
| | | Portion of agriculture area of holdings with mixed crops - livestock | agriculture area / km2 | | | | | | | | |
| Demo-graphy | Population | Population (23) | number | 2014-2023 | annually | NUTS 3 | m(1997-2015) | 2014 | m(2014 -16) | m(2014 -18) | m(2014-20) |
| | | Population in Spain(24) | number | 1997-2015 | >annually | NUTS 3 | m(2000, 2005, 2010, 2015) | | | | |

| | | | | | | | | | | |
|---|---|---|---|---|---|---|---|---|---|---|
| | Population density(25) | number / km2 | 2000, 2005, 2010, 2015, 2020 | snapshots | 30 arcsec | | m(2000, 2005, 2010) | m(2010 ,2015) | m(2010 ,2015) | m(2010,20 15,2020) |
| Demography (26) | Age dependency ratio, 1st variant (population 0 to 14 years and 65 years or over to population 15 to 64 years) | % | 2014-2023 | annually | NUTS 3 | m2014-15 | 2014 | m2014-16 | m(2014 -18) | m(2014-20) |
| | Age dependency ratio, 2nd variant (population 0 to 19 years and 60 years or over to population 20 to 59 years) | % | | | | | | | | |
| | Age dependency ratio, 3rd variant (population 0 to 19 years and 65 years or over to population 20 to 64 years) | % | | | | | | | | |
| | Age dependency ratio, 4th variant (population 0 to 24 years | % | | | | | | | | |

| | |
|---|---|
| and 65 years or over to population 25 to 64 years) | |
| Median age of population - females | year |
| Median age of population | year |
| Median age of population - males | year |
| Old-age dependency ratio 1st variant (population 65 years or over to population 15 to 64 years) | % |
| Old-age dependency ratio 2nd variant (population 60 years or over to population 20 to 59 years) | % |
| Old-age dependency ratio 3rd variant (population 65 years or over to population | % |

| | |
|---|---|
| 20 to 64 years) | |
| Old-age dependency ratio 4th variant (population 65 years or over to population 25 to 64 years) | % |
| Women per 100 men | % |
| Proportion of population aged 0-14 years | % |
| Proportion of population aged 0-18 years | % |
| Proportion of population aged 0-19 years | % |
| Proportion of population aged 0-4 years | % |
| Proportion of population aged 10-14 years | % |
| Proportion of population aged 15-19 years | % |

| | |
|---|---|
| Proportion of population aged 15-24 years | % |
| Proportion of population aged 15-29 years | % |
| Proportion of population aged 15-74 years | % |
| Proportion of population aged 20-24 years | % |
| Proportion of population aged 20-39 years | % |
| Proportion of population aged 20-64 years | % |
| Proportion of population aged 25-29 years | % |
| Proportion of population aged 25-44 years | % |
| Proportion of population aged 25-49 years | % |
| Proportion of population | % |

| | |
|---|---|
| aged 30-34 years | |
| Proportion of population aged 35-39 years | % |
| Proportion of population aged 40-44 years | % |
| Proportion of population aged 40-59 years | % |
| Proportion of population aged 45-49 years | % |
| Proportion of population aged 45-64 years | % |
| Proportion of population aged 50-54 years | % |
| Proportion of population aged 50-64 years | % |
| Proportion of population aged 55-59 years | % |
| Proportion of population aged 5-9 years | % |

| | |
|---|---|
| Proportion of population aged 60-64 years | % |
| Proportion of population aged 60-79 years | % |
| Proportion of population aged 60 years and more | % |
| Proportion of population aged 65-69 years | % |
| Proportion of population aged 65-79 years | % |
| Proportion of population aged 65 years and more | % |
| Proportion of population aged 70-74 years | % |
| Proportion of population aged 75-79 years | % |
| Proportion of population aged 75 years and more | % |

| | |
|---|---|
| Proportion of population aged 80-84 years | % |
| Proportion of population aged 80 years and more | % |
| Proportion of population aged 85 years and more | % |
| Proportion of population aged 100 years and more | % |
| Young-age dependency ratio 1st variant (population 0 to 14 years to population 15 to 64 years) | % |
| Young-age dependency ratio 2nd variant (population 0 to 19 years to population 20 to 59 years) | % |
| Young-age dependency ratio 3rd variant (population 0 | % |

| | | | | | | | | | | | |
|---|---|---|---|---|---|---|---|---|---|---|---|
| | | to 19 years to population 20 to 64 years) | | | | | | | | | |
| | | Young-age dependency ratio 4th variant (population 0 to 24 years to population 25 to 64 years) | % | | | | | | | | |
| Education (27) | Education | Population by educational attainment level | % | 2004-2023 | annually | NUTS 2 | m2004-15 | m(2004-2012) | m2011-16 | m(2009 -18) | m(2010-20) |
| Human footprint index(28) | Human footprint index | human footprint index | index | 2000-2022 | annually | 1km | m2000-15 | m(2000-2012) | m2011-16 | m(2009 -18) | m(2010-20) |
| Housing | Housing | Conventional dwelling(29) | number | 2011 | snapshot | NUTS 3 | 2011 | 2011 | 2011 | 2011 | 2011 |
| | | Population by housing arrangement (30) | number | 2011 | snapshot | NUTS 2 | 2011 | 2011 | 2011 | 2011 | 2011 |
| Inequality (31) | Inequality | quantile ratio S20/S80 | index | 2003-2023 | annually | NUTS 0-2 | 2021 | | m2011-16 | | 2021 |
| Income & Poverty | Income(32) | Disposable income of private households | Million purchasing power standards | 2011-2022 | annually | NUTS 2 | m2011-15 | m(2011-12) | m2011-16 | m(2011 -18) | m(2011-20) |
| | Poverty | At-risk-of-poverty rate (33) | % | 2003-2023 | annually | NUTS 2 | m2004-15 | m(2022-23) | m2011-16 | 2018 | m(2018-20) |
| | | Persons at risk of poverty or | % | 2014-2023 | annually | NUTS 2 | 2015 | m(2022-23) | m2015-16 | 2018 | m(2014-20) |

| | | | | | | | | | | | |
|---|---|---|---|---|---|---|---|---|---|---|---|
| | | social exclusion (34) | | | | | | | | | |
| Occupation | Unemploy-ment rate(35) | Unemploy-ment rate | % | 2012-2023 | annually | NUTS 2 | m2012-15 | 2012 | m2012-16 | m(2012 -18) | m(2012-20) |
| | Occupation (36) | Thousand employed persons | Thousand persons | 1995-2022 | annually | NUTS 3 | m2000-15 | m(2000-12) | m2011-16 | m(2009 -18) | m(2010-20) |
| Tourism | Tourism | nights private by origin region(37) | nights | 2018-2023 | annually | NUTS 2 | | | | | m(2018-20) |
| | | nights private by origin (DOM/FOR) (38) | nights | 2018-2023 | annually | NUTS 3 | | | | | m(2018-20) |
| | | nights by origin (DOM/FOR) (39) | nights / thousand inhabitants | 2020-2023 | annually | NUTS 3 | | | | | 2020 |
| | | | nights / km2 | | | | | | | | 2020 |
| | | total nights (39) | nights | | | | | | | | 2020 |
| | | Portion of DOM/FOR tourist nights (39) | % | | | | | | | | 2020 |
| Healthcare (40) | Healthcare capacity | number of beds | beds | 2020 | snapshot | NUTS 3 | 2020 | | 2020 | | |
| | | number of beds per inhabitant | beds / inhabitant | | | | 2020 | | 2020 | | |
| | | number of beds per area | beds / km2 | | | | 2020 | | 2020 | | |

| | | | | | | | | | | |
|---|---|---|---|---|---|---|---|---|---|---|
| | | | number of beds in public/private healthcare providers | | | | | 2020 | | |
| | | | number of beds per facility type group | | | | | 2020 | | 2020 |
| Human immuno-suppressive condition | Human immuno-suppressive condition (41) | HIV | prevalence of HIV day cases | cases / 100,1000 inhabitants | 2000-2021 | annually | NUTS 0-2 | m(2000-15) | m(2000-12) | 2011-2016 |
| | | | prevalence of HIV in-patients | cases / 100,1000 inhabitants | 2000-2021 | annually | NUTS 0-2 | m(2000-15) | m(2000-12) | 2011-2016 |
| | | | prevalence of HIV all cases | cases / 100,1000 inhabitants | 2000-2021 | annually | NUTS 0-2 | m(2000-15) | m(2000-12) | 2011-2016 |
| | | | 3 year average standardized death rate of HIV | % | 1996-2010 | annually | NUTS 0-2 | m(1996-2010) | m(1999-2010) | 2010 |
| | | Cancer | prevalence of cancer day cases | cases / 100,1000 inhabitants | 2000-2021 | annually | NUTS 0-2 | m(2000-15) | m(2000-12) | 2011-2016 |
| | | | prevalence of cancer in-patients | cases / 100,1000 inhabitants | 2000-2021 | annually | NUTS 0-2 | m(2000-15) | m(2000-12) | 2011-2016 |
| | | | prevalence of cancer | cases / 100,1000 inhabitants | 2000-2021 | annually | NUTS 0-2 | m(2000-15) | m(2000-12) | 2011-2016 |
| | | | 3 year average standardized death rate of cancer | % | 1996-2010 | annually | NUTS 2 | m(1997-2010) | m(1999-2010) | 2010 |

| | | | | | | | | | | | | |
|---|---|---|---|---|---|---|---|---|---|---|---|---|
| Biodiversity | Local birds(42) | Birds | Relative change in the number of bird species | % | 1900-2015 | decadal, 5 years | 1 degree | 2000, 2010, 2015 | 2000, 2010, 2015 | 2010, 2015 | 2010, 2015 | 2010, 2015 |
| | | | Absolute change in the number of bird species | number | | | | | | | | |
| | | Forest birds | Relative change in the number of forest bird species | % | | | | | | | | |
| | | | Absolute change in the number of forest bird species | number | | | | | | | | |
| | | Non-forest birds | Relative change in the number of non-forest bird species | % | | | | | | | | |
| | | | Absolute change in the number of non-forest bird species | number | | | | | | | | |
| | Birds(43) | Birds | Species richness of birds | number | 1900-2050 | 3 snapshots: 1900, 2015, 2050 | 1 degree | 2015 | 2015 | 2015 | 2015 | 2015 |
| | | | Species richness change of birds | % | | | | | | | | |
| | | | Weighted species richness | % | | | | | | | | |

| | | | | | | | | | | | |
|---|---|---|---|---|---|---|---|---|---|---|---|
| | | change of birds | | | | | | | | | |
| | Forest birds | Species richness of forest birds | number | | | | | | | | |
| | | Species richness change of forest birds | % | | | | | | | | |
| | | Weighted species richness change of forest birds | % | | | | | | | | |
| | Non-forest birds | Species richness of non-forest birds | number | | | | | | | | |
| | | Species richness change of non-forest birds | % | | | | | | | | |
| | | Weighted species richness change of non-forest birds | % | | | | | | | | |
| Vertebrates (44) | Vertebrates | Species richness of all vertebrates | number | 1900-2050 | 3 snapshots: 1900, 2015, 2050 | 1 degree | 2015 | 2015 | 2015 | 2015 | 2015 |
| | | Species richness change of all vertebrates | % | | | | | | | | |
| | | Weighted species richness | % | | | | | | | | |

| | | | | | | | | | | | | |
|---|---|---|---|---|---|---|---|---|---|---|---|---|
| | | | change of all vertebrates | | | | | | | | | |
| | | | Intactness of all vertebrates | - | | | | | | | | |
| | Mammals (45) | Mammals | Species richness of mammals | number | 1900-2050 | 3 snapshots: 1900, 2015, 2050 | 1 degree | 2015 | 2015 | 2015 | 2015 | 2015 |
| | | | Species richness change of mammals | % | | | | | | | | |
| | | | Weighted species richness change of mammals | % | | | | | | | | |
| Land use | Land use harmoniza-tion 2 (46) | Land use | C3 annual crops | fraction of grid cell | 1950-2019 | annually | 0.25 degree | 1997-2015 | 1999-2012 | 2011-2016 | 2009-2018 | 2010-2019 |
| | | | C3 nitrogen-fixing crops | fraction of grid cell | 1950-2019 | annually | | | | | | |
| | | | C3 perennial crops | fraction of grid cell | 1950-2019 | annually | | | | | | |
| | | | C4 annual crops | fraction of grid cell | 1950-2019 | annually | | | | | | |
| | | | C4 perennial crops | fraction of grid cell | 1950-2019 | annually | | | | | | |
| | | | Managed pasture | fraction of grid cell | 1950-2019 | annually | | | | | | |
| | | | Forested primary land | fraction of grid cell | 1950-2019 | annually | | | | | | |
| | | | Non-forested primary land | fraction of grid cell | 1950-2019 | annually | | | | | | |
| | | | Rangeland | fraction of grid cell | 1950-2019 | annually | | | | | | |

| | | | | | | | | | | | | |
|---|---|---|---|---|---|---|---|---|---|---|---|---|
| | | | Potentially forested secondary land | fraction of grid cell | 1950-2019 | annually | | | | | | |
| | | | Potentially non-forested secondary land | fraction of grid cell | 1950-2019 | annually | | | | | | |
| | | | Secondary mean age | years | 1950-2019 | annually | | | | | | |
| | | | Secondary mean biomass carbon density | kg m-2 | 1950-2019 | annually | | | | | | |
| | | | Urban land | fraction of grid cell | 1950-2019 | annually | | | | | | |
| Bio-climate | Climatology (47) | Temperature | | | 1979-2018 | 40 years mean | 0.5 degree | 1979-2018 | 1979-2018 | 1979-2018 | 1979-2018 | 1979-2018 |
| | | Growing-degree-days | | | | | | | | | | |
| | | Growing season | | | | | | | | | | |
| | | Summer-days | | | | | | | | | | |
| | | Frost days | | | | | | | | | | |
| | | Surface sensitive heat flux | | | | | | | | | | |
| | | Precipitation | | | | | | | | | | |
| | | Aridity | | | | | | | | | | |
| | | Dry-days | | | | | | | | | | |
| | | Dry-spells | | | | | | | | | | |
| | | Evaporative fraction | | | | | | | | | | |

| |
|---|
| Potential evaporation |
| Water vapor pressure |
| Surface latent heat flux |
| Soil moisture |
| Wind speed |
| Water vapor pressure |
| Meridional wind speed |
| Zonal wind speed |

Columns nine to 13 show the used time periods of the predictor variables to best match with the time span of the dependent variables.
SF = sand fly
CanL = canine leishmaniasis
FelL = feline leishmaniasis
DOM = domestic
FOR = foreign
HIV = human immunodeficiency virus
m() = the mean of
NA = metadata unavailable
grey cells = data unavailable

#### S3. Model formulation

This document was constructed based on Moraga (2019, 2014) (48,49) and Zuur & Ieno (2017) (50).

When expected incidence could be calculated given data, let $Y_i$ and $E_i$ be the observed and expected number of leishmaniasis, respectively, and let $\theta_i$ be the relative risk for NUTS3 $i = 1, \ldots, n$. The model is specified as follows:

$$Y_i|\theta_i \sim Poisson\ (E_i \times \theta_i),\ i = 1, \ldots, n, \quad (1)$$

$$\log(\theta_i) = \beta_0 + u_i + \varepsilon \quad (2)$$

$$\log(\theta_i) = \beta_0 + S(x_i) + \varepsilon \quad (3)$$

$$\log(\theta_i) = \beta_0 + \beta_1 \times var_i + \varepsilon \quad (4)$$

$$\log(\theta_i) = \beta_0 + f(var_i) + \varepsilon \quad (5)$$

$$\log(\theta_i) = \beta_0 + \beta_1 \times var_i + u_i + \varepsilon \quad (6)$$

$$\log(\theta_i) = \beta_0 + f(var_i) + u_i + \varepsilon \quad (7)$$

$$\log(\theta_i) = \beta_0 + \beta_1 \times var_i + S(x_i) + \varepsilon \quad (8)$$

$$\log(\theta_i) = \beta_0 + f(var_i) + S(x_i) + \varepsilon \quad (9)$$

where, $\beta_0$ is the intercept and $\beta_1$ is the coefficient of the covariate $var_i$. $f(\cdot)$ is smooth function of the covariate $var_i$ follows a random walk of order 2. $u_i$ is a conditionally autoregressive model, $u_i|\mathbf{u}_{-i} \sim N(\bar{u}_{\delta_i} \frac{1}{\tau_u n_{\delta_i}})$. $S(\cdot)$ is a spatial random effect that follows a zero-mean Gaussian process with Matérn covariance function

$$\mathrm{Cov}(S(x_i), S(x_j)) = \frac{\sigma^2}{2^{v-1}\Gamma(v)} \left(\kappa||x_i - x_j||\right)^v K_v(\kappa||x_i - x_j||), \quad (10)$$

Where $K_v(\cdot)$ is the modified Bessel function of second kind and order $v > 0$. $v$ is the smoothness parameter, $\sigma^2$ denotes the variance, and $\kappa > 0$ is related to the practical range $\rho = \sqrt{8v}/\kappa$ which is the distance at which the spatial correlation is close to 0.1.

Formular (2) – (9) are null model of neighbor structure (m0_nb), null model of distance-based spatial random effect (m0_spde), generalized linear model (GLM), generalized additive model (GAM), generalized linear model with neighbor structure (spGLM_nb), generalized linear model with distance-based spatial random effect (spGLM_spde), generalized additive model with neighbor structure (spGAM_nb), generalized additive model with distance-based spatial random effect (spGAM_spde).

Extending the one variable models with the second variable and the interaction between the two variables, formula (4) – (9) become:

$$\log(\theta_i) = \beta_0 + \beta_1 \times var_{1,i} + \beta_2 \times var_{2,i} + \beta_3 \times var_{1,i} \times var_{2,i} + \varepsilon \quad (11)$$

$$\log(\theta_i) = \beta_0 + f(var_{1,i}) + f(var_{2,i}) + f(var_{1,i} \times var_{2,i}) + \varepsilon \quad (12)$$

$$\log(\theta_i) = \beta_0 + \beta_1 \times var_{1,i} + \beta_2 \times var_{2,i} + \beta_3 \times var_{1,i} \times var_{2,i} + u_i + \varepsilon \quad (13)$$

$$\log(\theta_i) = \beta_0 + f(var_{1,i}) + f(var_{2,i}) + f(var_{1,i} \times var_{2,i}) + u_i + \varepsilon \quad (14)$$

$$\log(\theta_i) = \beta_0 + \beta_1 \times var_{1,i} + \beta_2 \times var_{2,i} + \beta_3 \times var_{1,i} \times var_{2,i} + S(x_i) + \varepsilon \quad (15)$$

$$\log(\theta_i) = \beta_0 + f(var_{1,i}) + f(var_{2,i}) + f(var_{1,i} \times var_{2,i}) + S(x_i) + \varepsilon \quad (16)$$

Where and $\beta_2$ is the coefficient of the covariate $var_{2,i}$, and $\beta_3$ is the coefficient of the interaction term between covariates $var_{1,i}$ and $var_{2,i}$.

When expected incidence could not be calculated given data, formula (1) - (9) and (11)-(16) become:

$$Y_i \sim Poisson\ (E_i),\ i = 1, \ldots, n, \quad (17)$$

$$\log(E_I) = \beta_0 + u_i + \varepsilon \quad (18)$$

$$\log(E_i) = \beta_0 + S(x_i) + \varepsilon \quad (19)$$

$$\log(E_i) = \beta_0 + \beta_i \times var_i + \varepsilon \quad (20)$$

$$\log(E_i) = \beta_0 + f(var_i) + \varepsilon \quad (21)$$

$$\log(E_i) = \beta_0 + \beta_i \times var_i + u_i + \varepsilon \quad (22)$$

$$\log(E_i) = \beta_0 + f(var_i) + u_i + \varepsilon \quad (23)$$

$$\log(E_i) = \beta_0 + \beta_i \times var_i + S(x_i) + \varepsilon \quad (24)$$

$$\log(E_i) = \beta_0 + f(var_i) + S(x_i) + \varepsilon \quad (25)$$

$$\log(E_i) = \beta_0 + \beta_1 \times var_{1,i} + \beta_2 \times var_{2,i} + \beta_3 \times var_{1,i} \times var_{2,i} + \varepsilon \quad (26)$$

$$\log(E_i) = \beta_0 + f(var_{1,i}) + f(var_{2,i}) + f(var_{1,i} \times var_{2,i}) + \varepsilon \quad (27)$$

$$\log(E_i) = \beta_0 + \beta_1 \times var_{1,i} + \beta_2 \times var_{2,i} + \beta_3 \times var_{1,i} \times var_{2,i} + u_i + \varepsilon \quad (28)$$

$$\log(E_i) = \beta_0 + f(var_{1,i}) + f(var_{2,i}) + f(var_{1,i} \times var_{2,i}) + u_i + \varepsilon \quad (29)$$

$$\log(E_i) = \beta_0 + \beta_1 \times var_{1,i} + \beta_2 \times var_{2,i} + \beta_3 \times var_{1,i} \times var_{2,i} + S(x_i) + \varepsilon \quad (30)$$

$$\log(E_i) = \beta_0 + f(var_{1,i}) + f(var_{2,i}) + f(var_{1,i} \times var_{2,i}) + S(x_i) + \varepsilon \quad (31)$$

For dataset with excessive zero's, hurdle model or zero-altered Poisson (ZAP) was employed. ZAP consists of a Bernoulli regression as classifier and a zero-truncated Poisson (ZTP) for the regions where the response variable is not zero.

$$Y_i \sim ZAP\ (E_i, \pi_i),\ i = 1, \ldots, n, \quad (32)$$

Where $\pi$ is the probability of the presence of leishmaniasis. Formulas (18)-(31) can be applied to ZTP. The Bernoulli model is defined as follows:

$$\text{logit}(\pi_i) = \gamma_0 + u_i + \varepsilon \quad (33)$$

$$\text{logit}(\pi_i) = \gamma_0 + S(x_i) + \varepsilon \quad (34)$$

$$\text{logit}(\pi_i) = \gamma_0 + \gamma_i \times var_i + \varepsilon \quad (35)$$

$$\text{logit}(\pi_i) = \gamma_0 + f(var_i) + \varepsilon \quad (36)$$

$$\text{logit}(\pi_i) = \gamma_0 + \gamma_i \times var_i + u_i + \varepsilon \quad (37)$$

$$\text{logit}(\pi_i) = \gamma_0 + f(var_i) + u_i + \varepsilon \quad (38)$$

$$\text{logit}(\pi_i) = \gamma_0 + \gamma_i \times var_i + S(x_i) + \varepsilon \quad (39)$$

$$\text{logit}(\pi_i) = \gamma_0 + f(var_i) + S(x_i) + \varepsilon \quad (40)$$

$$\text{logit}(\pi_i) = \gamma_0 + \gamma_1 \times var_{1,i} + \gamma_2 \times var_{2,i} + \gamma_3 \times var_{1,i} \times var_{2,i} + \varepsilon \quad (41)$$

$$\text{logit}(\pi_i) = \gamma_0 + f(var_{1,i}) + f(var_{2,i}) + f(var_{1,i} \times var_{2,i}) + \varepsilon \quad (42)$$

$$\text{logit}(\pi_i) = \gamma_0 + \gamma_1 \times var_{1,i} + \gamma_2 \times var_{2,i} + \gamma_3 \times var_{1,i} \times var_{2,i} + u_i + \varepsilon \quad (43)$$

$$\text{logit}(\pi_i) = \gamma_0 + f(var_{1,i}) + f(var_{2,i}) + f(var_{1,i} \times var_{2,i}) + u_i + \varepsilon \quad (44)$$

$$\text{logit}(\pi_i) = \gamma_0 + \gamma_1 \times var_{1,i} + \gamma_2 \times var_{2,i} + \gamma_3 \times var_{1,i} \times var_{2,i} + S(x_i) + \varepsilon \quad (45)$$

$$\text{logit}(\pi_i) = \gamma_0 + f(var_{1,i}) + f(var_{2,i}) + f(var_{1,i} \times var_{2,i}) + S(x_i) + \varepsilon \quad (46)$$

where, $\gamma_0$ is the intercept and $\gamma_1$ is the coefficient of the covariate $var_i$ and $var_{1,i}$, $\gamma_2$ is the coefficient of the covariate $var_{2,i}$, and $\gamma_3$ is the coefficient of the interaction term between covariates $var_{1,i}$ and $var_{2,i}$. The covariates $var_i$, $var_{1,i}$ and $var_{2,i}$ in the Bernoulli model and in ZTP can differ.

When the number of test is fixed, conditional on the true prevalence $P(x_i)$ at location $x_i$, $i = 1, \ldots., n$, the number of positive results $Y_i$ out of $N_i$ people sampled follows a binomial distribution:

$$Y_i | P(x_i) \sim Binomial\,(N_i, P(x_i)),\ i = 1, \ldots., n, \quad (47)$$

$$\text{logit}(P(x_i)) = \beta_0 + u_i + \varepsilon \quad (48)$$

$$\text{logit}(P(x_i)) = \beta_0 + S(x_i) + \varepsilon \quad (49)$$

$$\text{logit}(P(x_i)) = \beta_0 + \beta_i \times var_i + \varepsilon \quad (50)$$

$$\text{logit}(P(x_i)) = \beta_0 + f(var_i) + \varepsilon \quad (51)$$

$$\text{logit}(P(x_i)) = \beta_0 + \beta_i \times var_i + u_i + \varepsilon \quad (52)$$

$$\text{logit}(P(x_i)) = \beta_0 + f(var_i) + u_i + \varepsilon \quad (53)$$

$$\text{logit}(P(x_i)) = \beta_0 + \beta_i \times var_i + S(x_i) + \varepsilon \quad (54)$$

$$\text{logit}(P(x_i)) = \beta_0 + f(var_i) + S(x_i) + \varepsilon \quad (55)$$

$$\text{logit}(P(x_i)) = \beta_0 + \beta_1 \times var_{1,i} + \beta_2 \times var_{2,i} + \beta_3 \times var_{1,i} \times var_{2,i} + \varepsilon \quad (56)$$

$$\text{logit}(P(x_i)) = \beta_0 + f(var_{1,i}) + f(var_{2,i}) + f(var_{1,i} \times var_{2,i}) + \varepsilon \quad (57)$$

$$\text{logit}(P(x_i)) = \beta_0 + \beta_1 \times var_{1,i} + \beta_2 \times var_{2,i} + \beta_3 \times var_{1,i} \times var_{2,i} + u_i + \varepsilon \quad (58)$$

$$\text{logit}(P(x_i)) = \beta_0 + f(var_{1,i}) + f(var_{2,i}) + f(var_{1,i} \times var_{2,i}) + u_i + \varepsilon \quad (59)$$

$$\text{logit}(P(x_i)) = \beta_0 + \beta_1 \times var_{1,i} + \beta_2 \times var_{2,i} + \beta_3 \times var_{1,i} \times var_{2,i} + S(x_i) + \varepsilon \quad (60)$$

$$\text{logit}(P(x_i)) = \beta_0 + f(var_{1,i}) + f(var_{2,i}) + f(var_{1,i} \times var_{2,i}) + S(x_i) + \varepsilon \quad (61)$$

27. Eurostat. Eurostat [Internet]. [cited 2025 Jun 2]. Population by educational attainment level, sex, age, country of birth and NUTS 2 region (%) (edat_lfs_9917). Available from:

https://ec.europa.eu/eurostat/databrowser/view/edat_lfs_9917/default/table?lang=en doi:https://doi.org/10.2908/EDAT_LFS_9917

37. Eurostat. Eurostat [Internet]. [cited 2024 Oct 18]. Guest nights spent at short-stay accommodation offered via collaborative economy platforms by NUTS 1 and NUTS 2 region, by world region of residence of the guest - experimental statistics (tour_ce_oaw). Available from:

https://ec.europa.eu/eurostat/databrowser/view/tour_ce_oaw/default/table?lang=en doi:https://doi.org/10.2908/TOUR_CE_OAW

49. Moraga Paula. Spatial statistics for data science : theory and practice with R. CRC Press; 2024.

50. Zuur AF, Ieno EN. GAM and Zero-Inflated Models. II. 2017.
